# Disparities in Quality of Healthcare between Asian and White Children with Special Health Care Needs from 2016-2020

**DOI:** 10.1101/2025.08.18.25333922

**Authors:** Kayla K. Van, Chin-Chih Chen, Esther Son, Ruth C. Brown

**Affiliations:** Department of Psychology, Virginia Commonwealth University, 806 W. Franklin St., Box 842018, Richmond, Virginia 23284-2018; Department of Counseling and Special Education, Virginia Commonwealth University, 1015 West Main Street, Box 842020, Richmond, VA 23284-2020; Department of Social Work, College of Staten Island, City University of New York, 2800 Victory Blvd., Staten Island, NY 10314, Office: 2A, 201-C; Department of Psychiatry, Virginia Commonwealth University, 800 East Leigh St., Suite 101, Richmond, VA 23219-1534

**Keywords:** Asian, children, health care quality, special healthcare needs, disability, health care disparities

## Abstract

**Background:** Approximately one in five families in the United States have a child with a special healthcare need (CSHCN). Previous research has shown significant inequalities in health care quality, access, satisfaction, and outcomes of Asian compared to White individuals. There is limited research currently available exploring disparities in health care quality at the intersection of race and CSHCN complexity for Asian versus White children.

**Objective:** The goal of this study is to understand how race, household language, poverty, family structure, and complexity of healthcare needs affect the quality of health care received by Asian and White CSHCNs.

**Method:** Data was obtained from the 2016-2020 National Survey of Children’s Health. Five multivariate ordinal regression analyses were performed to explore the predictors of parents’ perceptions of healthcare quality across five domains: time spent with the child, doctor listening to parent concerns, doctor being sensitive to family values/customs, doctor providing needed information, and doctor partnering with the family in their child’s care.

**Results:** Results show that Asian parents with CHSCNs report lower quality care than White parents with CSHCNs. However, quality of care dropped for high complexity CSHCN regardless of race.

Significant covariates included language barriers, socioeconomic factors, and cultural differences.

**Conclusion:** Asian CSHCNs experience disparities in healthcare quality, access, and utilization compared to their White counterparts. The results from this study suggest the need to examine subgroup differences and develop effective intervention strategies to ensure equitable care for everyone, especially CSHCN with high complex needs.

---

One in five families in the United States has a child with a special healthcare need (US; CSHCN; Jolles et al., 2018). According to the Maternal and Child Health Bureau (MCHB), classifying CSHCNs includes children who are at an elevated risk and have a chronic physical, developmental, behavioral, or emotional condition and require healthcare services more often than other children generally do (Van Dyck et al., 2004). This group of children can include those with asthma, autism, ADHD, anxiety, and depression. The prevalence of special healthcare needs was highest among boys, children ages 6-12, and children in low-income families (Van Dyck et al., 2004). Health outcomes of CSHCNs are worse compared to children without special healthcare needs (Bramlett et al., 2009; Cheak-Zamora & Thullen, 2017; Houtrow et al., 2011).

The population of CSHCNs has continued to rise within the past decade, creating challenges to reform healthcare with its goals to enhance population health, improve patients’ experience and quality of care, and reduce costs (Strickland et al., 2015). However, most parents with CSHCN report having personal challenges in health care and related systems (Hill et al., 2008; Jolles et al., 2018). These obstacles can include minimal sources of relevant information, lack of proper insurance coverage for CSHCNs, and a shortage of physicians prepared to treat CSHCNs due to their limited understanding of the specific healthcare needs of these individuals (Hill et al., 2008; Jolles et al., 2018; Magaña et al., 2012). About 18% of CSHCNs reported having an unmet need for a specific health care service, such as primary and specialty care services, dental care (8.1%), and mental health care (1.2%) (Van Dyck et al., 2004; Hill et al., 2008). In addition to having an unmet need for health care services, 22% of CSHCNs had trouble acquiring a referral they needed (Van Dyck et al., 2004).

Race and ethnicity play a significant role in healthcare utilization rates in the US and are recognized as social determinants of health (Mehta et al., 2013). Even though awareness of racial and ethnic disparities in healthcare has grown, research on healthcare inequalities among children has declined (Flores & Tomany-Korman, 2008). It has been shown that minoritized CSHCNs experience significant inequalities in healthcare quality, access, satisfaction, and outcomes compared to White CSHCNs and those without disabilities (Son et al., 2018). Parental limited English proficiency is associated with poorer health outcomes among minoritized children, contributing as one of the social determinants of health (Eneriz-Wiemer et al., 2014).

Despite many studies concentrating on minority groups in the US, there is still a lack of research that explores the Asian population and their experiences. According to a past study that examined health status and disparities in health care access and utilization among children from the six largest Asian ethnic groups in California, they found that factors such as age, gender, poverty, citizenship and nativity status, health insurance, and parental marital and child health statuses were associated with health care access and utilization among Asian children compared to non-Hispanic White children (Yu et al., 2010). Cultural differences also contribute to the decrease in healthcare utilization in the US. Health problems, such as difficulty in accessing health services, are caused by the issue of adapting to a new culture, specifically for children with limited English proficiency (Yu et al., 2004). Asian immigrant parents of CSHCN are also less likely to visit doctors in the past year, possibly due to cultural and language barriers that stand between them and the healthcare provider (Javier et al., 2010).

Several studies have examined racial and ethnic disparities in healthcare quality using parent-reported measures. Ngui and Flores (2005) explored racial and ethnic disparities in healthcare among CSHCNs based on parent-reported responses in the National Survey of Children with Special Health Care Needs. Their study assessed parental satisfaction based on variables such as the amount of time doctors spent with the child and whether physicians carefully listened to parental concerns (Ngui and Flores, 2005). Results suggest that Black and Hispanic parents were more likely to be dissatisfied with care and to report issues with the convenience of service use compared to White parents (Ngui and Flores, 2005). Additionally, the severity of the child’s condition, lack of insurance, and insufficient family-centered care were heavily associated with dissatisfaction with care and difficulties with easy use of health care services (Ngui and Flores, 2005).

Similarly, Morgan et al. (2023) also found comparable results using the same measure in the National Survey of Children’s Health (NSCH) survey. The study explored disparities in family-centered care among children and youths with special health care needs in the US (Morgan et al., 2023). Utilizing the same variables as Ngui and Flores (2005) and Son et al. (2017), they also found that parents of children and youth with special healthcare needs who are Black or Hispanic, experiencing poverty, and lack health insurance coverage report acquiring lower quality family-centered care compared to White parents (Morgan et al., 2023). Furthermore, they discovered that language use, poverty, disability status, and the severity of the children’s disability also contributed to the disparities in obtaining family-centered care (Morgan et al., 2023).

Although several studies focus on the disparities in healthcare among minorities, very few studies specifically investigate the healthcare quality of Asian CSHCNs (Son et al., 2017). One study found that Asian parents of CSHCNs reported that their healthcare providers were less likely to give them the exact information they needed, to make them feel like a partner in their child’s care, and to be understanding of their family’s values and customs (Son et al., 2017). Further research is needed to better understand these gaps. Using data from the NSCH, this study examines how race, household language, poverty, family structure, and other factors influence the quality of health care received by Asian and White CSHCNs. This study aims to address the gap in understanding healthcare quality disparities among Asian CSHCNs, highlight the unique challenges faced by Asian families, and underscore the need for more culturally responsive healthcare practices to promote equity in pediatric care.

## Methods

### Participants

This study utilized publicly available and anonymized data from the NSCH from 2016-2020, provided by the Data Resource Center for Child & Adolescent Health (www.childhealthdata.org). As the research does not involve interaction with human participants or identifiable personal data, it is not considered human subjects research and is exempt from approval from the Institutional Review Board. The NSCH is funded through the MCHB and follows Title V National Performance Measures and National Outcome Measures. A total of 109,626 were included in the analyses. The sample is comprised of 93.90% (n = 102,938 non-Hispanic White and 6.10% (n = 6,688) non-Hispanic Asian children, ages 3-17.

### Measures

#### Dependent Variables

Healthcare quality was assessed using five criteria based on parents’ responses about their experiences with providers. “Time” assessed whether doctors spent enough time with their child. “Listen” evaluated whether providers listened carefully to parents’ concerns. “Sensitivity” measured whether physicians demonstrated sensitivity to the family’s values and customs. “Information” examined whether providers supplied information specific to parents’ concerns. “Partner” assessed whether doctors helped parents feel like partners in their child’s care. Responses were coded “Sometimes or Never,” “Usually,” or “Always,” with higher numbers reflecting higher quality of care. Missing responses and respondents who reported not having a medical visit in the last year were excluded from the analysis.

#### Independent Variable

Race was established as the primary independent variable, specifically non-Hispanic Asian and non-Hispanic White children. Non-Hispanic White children were used as a reference group.

#### Covariates

Covariates consist of diverse social and economic factors. Social variables included: *primary household language* (English and other than English), *parent nativity* (all parents born in the US (3rd or higher generation household); any parent born outside of the US (1st and 2nd generation household); other (child born in the US, parents are not listed)), and *family structure* (two parents, currently married; two parents, not currently married; other family type). Economic variables comprised the *household’s income level* (0-99% FPL; 100-199% FPL; 200-399% FPL; 400% FPL or greater) that the child lives in and the *adequacy of insurance coverage* (adequate and inadequate). Additionally, we examined the complexity of CSHCN (non-CSHCN, less complex, and more complex). The reference groups included the household language to be English; both parents born in the US; two parents, currently married; household income level at 400% FPL or greater; adequate insurance coverage; and non-CSHCN complexity.

### Data Analysis

Data analyses were conducted using R (version 4.4.2, running on Windows 10 x64). Parent-reported information on the five criteria of health care were used as outcomes. Household language, poverty level, family structure, parent nativity, insurance type, adequacy of current insurance, and complexity of CSHCN were included as main effect predictors. An interaction between race and CSHCN complexity was included. Five multivariate ordinal regressions, one for each healthcare quality outcome, were created using ‘clm()’ in the package ‘ordinal’.

## Results

### Descriptive Statistics

Data were drawn from a sample of 109,626 families, comprising 102,938 White, non-Hispanic families and 6,688 Asian, non-Hispanic families. As shown in Table 1, across all variables, chi-square tests indicated statistically significant differences between White and Asian families (ps < .001). Most White families (99%) reported speaking English at home, whereas only 64% of Asian families reported English as the primary household language. The two groups also differed in socioeconomic status: although approximately half of both White (48%) and Asian (53%) families were at or above 400% of the federal poverty level (FPL), a slightly higher proportion of Asian families (8.7%) than White families (7.1%) fell below 100% FPL. Regarding insurance coverage, the majority of families had adequate insurance (67% of White families, 69% of Asian families), but about one-third of each group reported inadequate coverage.

**Table 1.**
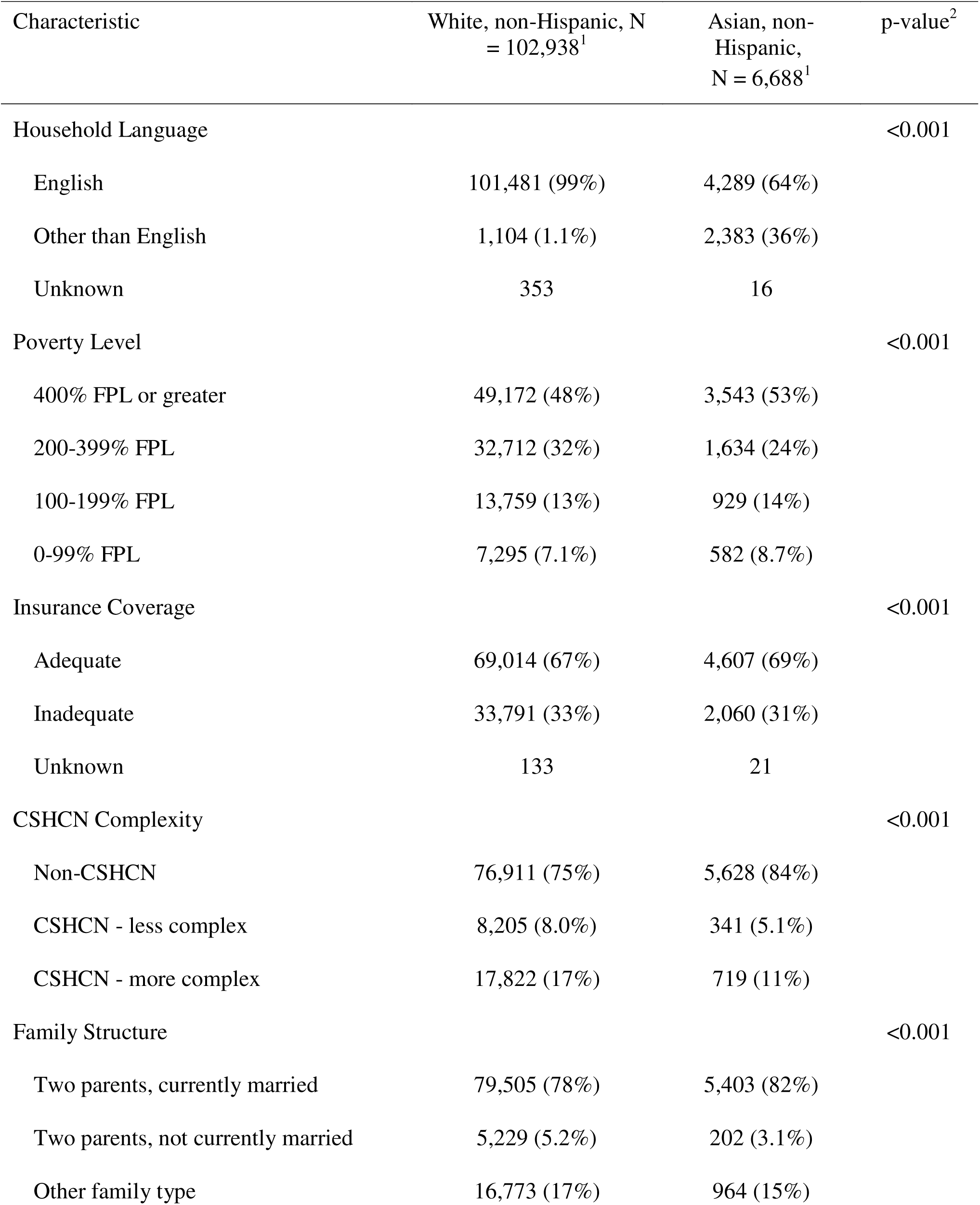
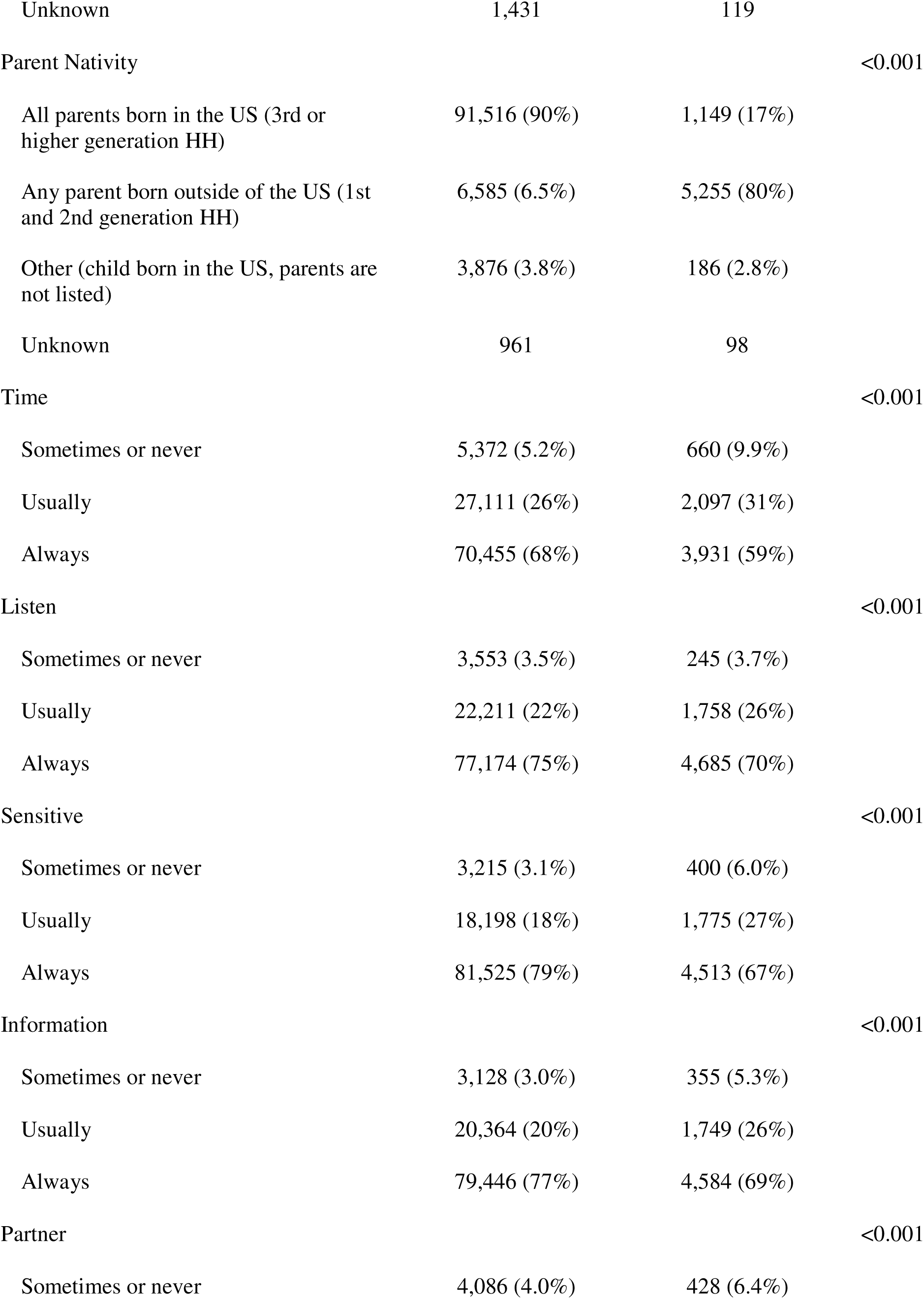
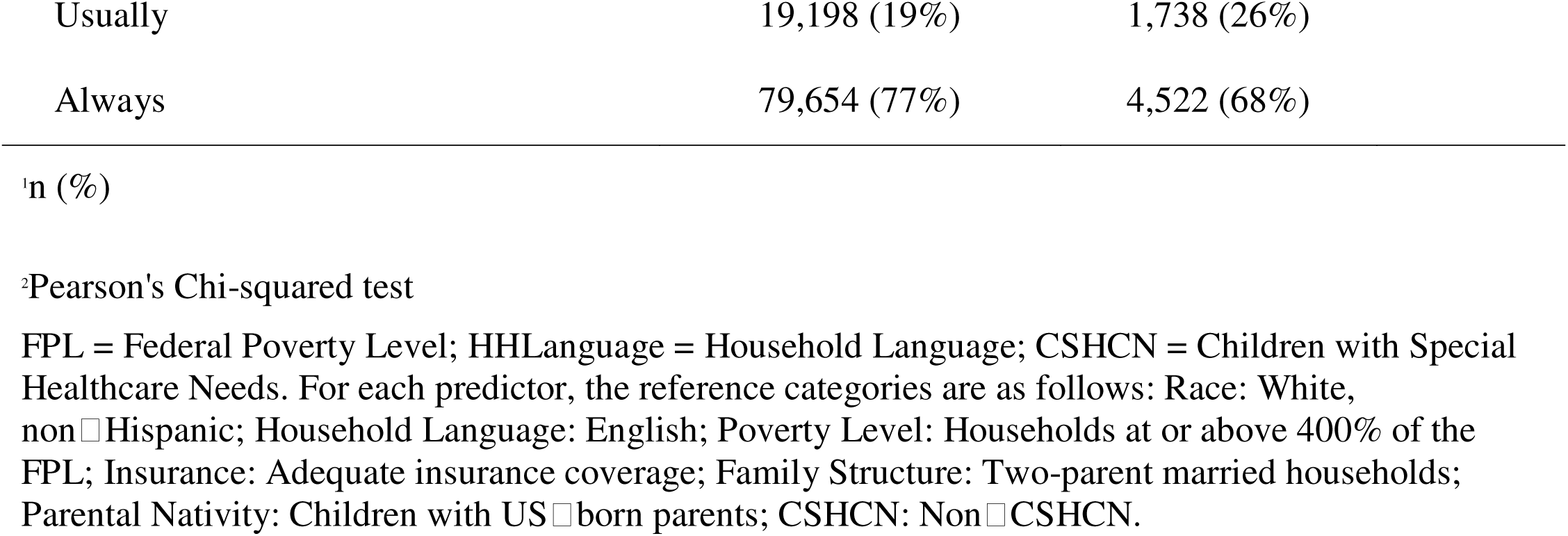
Demographic Characteristics by Race.

| Characteristic | White, non-Hispanic, N<br>= 102,938<sup>1</sup> | Asian, non-Hispanic,<br>N = 6,688<sup>1</sup> | p-value<sup>2</sup> |
| --- | --- | --- | --- |
| Household Language |  |  | <0.001 |
| English | 101,481 (99%) | 4,289 (64%) |  |
| Other than English | 1,104 (1.1%) | 2,383 (36%) |  |
| Unknown | 353 | 16 |  |
| Poverty Level |  |  | <0.001 |
| 400% FPL or greater | 49,172 (48%) | 3,543 (53%) |  |
| 200-399% FPL | 32,712 (32%) | 1,634 (24%) |  |
| 100-199% FPL | 13,759 (13%) | 929 (14%) |  |
| 0-99% FPL | 7,295 (7.1%) | 582 (8.7%) |  |
| Insurance Coverage |  |  | <0.001 |
| Adequate | 69,014 (67%) | 4,607 (69%) |  |
| Inadequate | 33,791 (33%) | 2,060 (31%) |  |
| Unknown | 133 | 21 |  |
| CSHCN Complexity |  |  | <0.001 |
| Non-CSHCN | 76,911 (75%) | 5,628 (84%) |  |
| CSHCN - less complex | 8,205 (8.0%) | 341 (5.1%) |  |
| CSHCN - more complex | 17,822 (17%) | 719 (11%) |  |
| Family Structure |  |  | <0.001 |
| Two parents, currently married | 79,505 (78%) | 5,403 (82%) |  |
| Two parents, not currently married | 5,229 (5.2%) | 202 (3.1%) |  |
| Other family type | 16,773 (17%) | 964 (15%) |  |
| Unknown | 1,431 | 119 |  |
| Parent Nativity |  |  | <0.001 |
| All parents born in the US (3rd or higher generation HH) | 91,516 (90%) | 1,149 (17%) |  |
| Any parent born outside of the US (1st and 2nd generation HH) | 6,585 (6.5%) | 5,255 (80%) |  |
| Other (child born in the US, parents are not listed) | 3,876 (3.8%) | 186 (2.8%) |  |
| Unknown | 961 | 98 |  |
| Time |  |  | <0.001 |
| Sometimes or never | 5,372 (5.2%) | 660 (9.9%) |  |
| Usually | 27,111 (26%) | 2,097 (31%) |  |
| Always | 70,455 (68%) | 3,931 (59%) |  |
| Listen |  |  | <0.001 |
| Sometimes or never | 3,553 (3.5%) | 245 (3.7%) |  |
| Usually | 22,211 (22%) | 1,758 (26%) |  |
| Always | 77,174 (75%) | 4,685 (70%) |  |
| Sensitive |  |  | <0.001 |
| Sometimes or never | 3,215 (3.1%) | 400 (6.0%) |  |
| Usually | 18,198 (18%) | 1,775 (27%) |  |
| Always | 81,525 (79%) | 4,513 (67%) |  |
| Information |  |  | <0.001 |
| Sometimes or never | 3,128 (3.0%) | 355 (5.3%) |  |
| Usually | 20,364 (20%) | 1,749 (26%) |  |
| Always | 79,446 (77%) | 4,584 (69%) |  |
| Partner |  |  | <0.001 |
| Sometimes or never | 4,086 (4.0%) | 428 (6.4%) |  |

|  |  |  |
| --- | --- | --- |
| Usually | 19,198 (19%) | 1,738 (26%) |
| Always | 79,654 (77%) | 4,522 (68%) |

<sup>a</sup>Pearson's Chi-squared test
FPL = Federal Poverty Level; HHLanguage = Household Language; CSHCN = Children with Special Healthcare Needs. For each predictor, the reference categories are as follows: Race: White, non-Hispanic; Household Language: English; Poverty Level: Households at or above 400% of the FPL; Insurance: Adequate insurance coverage; Family Structure: Two-parent married households; Parental Nativity: Children with US-born parents; CSHCN: Non-CSHCN.

CSHCNs were more prevalent among White families (25%) than Asian families (16%). Specifically, 17% of White families had a child with more complex needs, whereas 11% of Asian families had a child with more complex needs. Family structure varied as well; 78% of White families and 82% of Asian families consisted of two parents who were currently married, with smaller proportions in other family types. Parental nativity revealed a stark contrast: 90% of White families had all parents born in the US, whereas 80% of Asian families had at least one parent born outside the US.

### Racial Differences in Healthcare Quality

Parents’ reports of healthcare quality also differed by race/ethnicity (Table 1). For instance, 68% of White families felt their child’s doctor “Always” spent enough time with their child, compared with 59% of Asian families. Similar patterns emerged for other outcomes, such as whether doctors listened to parent concerns (75% vs. 70% reporting “Always”), showed sensitivity to family values/customs (79% vs. 67%), provided needed information (77% vs. 69%), and partnered with the family (77% vs. 68%). Chi-square tests indicated statistically significant differences across all healthcare quality measures.

### Multivariate Predictors of Healthcare Quality

Five separate ordinal logistic regression models were estimated to examine predictors of parents’ perceptions of healthcare quality across five domains: *Time spent with child*, *Doctor listening to parent concerns*, *Doctor being sensitive to family values/customs*, *Doctor providing needed information*, and *Doctor partnering with the family*. Table 2 presents the odds ratios (ORs) and standard errors (SEs) for each predictor in each model, along with overall fit indices (AIC, BIC, Log-Likelihood, McFadden R²). Figure 1 displays the corresponding predicted probabilities of reporting “Sometimes or Never,” “Usually,” or “Always,” plotted by Race/Ethnicity (White, Asian) and CSHCN complexity level (Non-CSHCN, Less Complex, More Complex).

**Figure 1.**
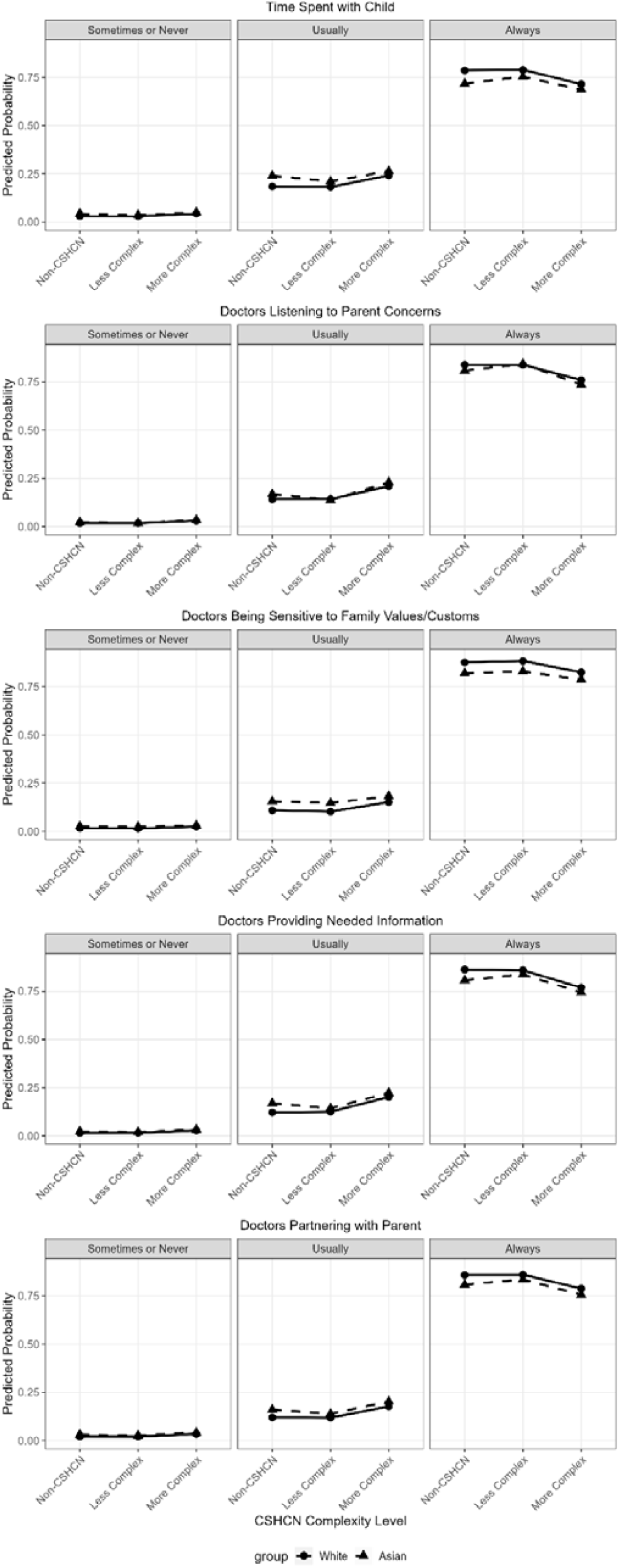
Predicted probabilities of parent-reported care quality outcomes by CSHCN Complexity and Race/Ethnicity, based on ordinal regression models. Outcomes include (from top to bottom): time spent with child, doctors listening to parent concerns, doctors being sensitive to family values/customs, doctors providing needed information, and doctors partnering with parent. Solid lines with circles represent White parents; dashed lines with triangles represent Asian parents. Shaded areas indicate 95% confidence intervals.

**Table 2.**
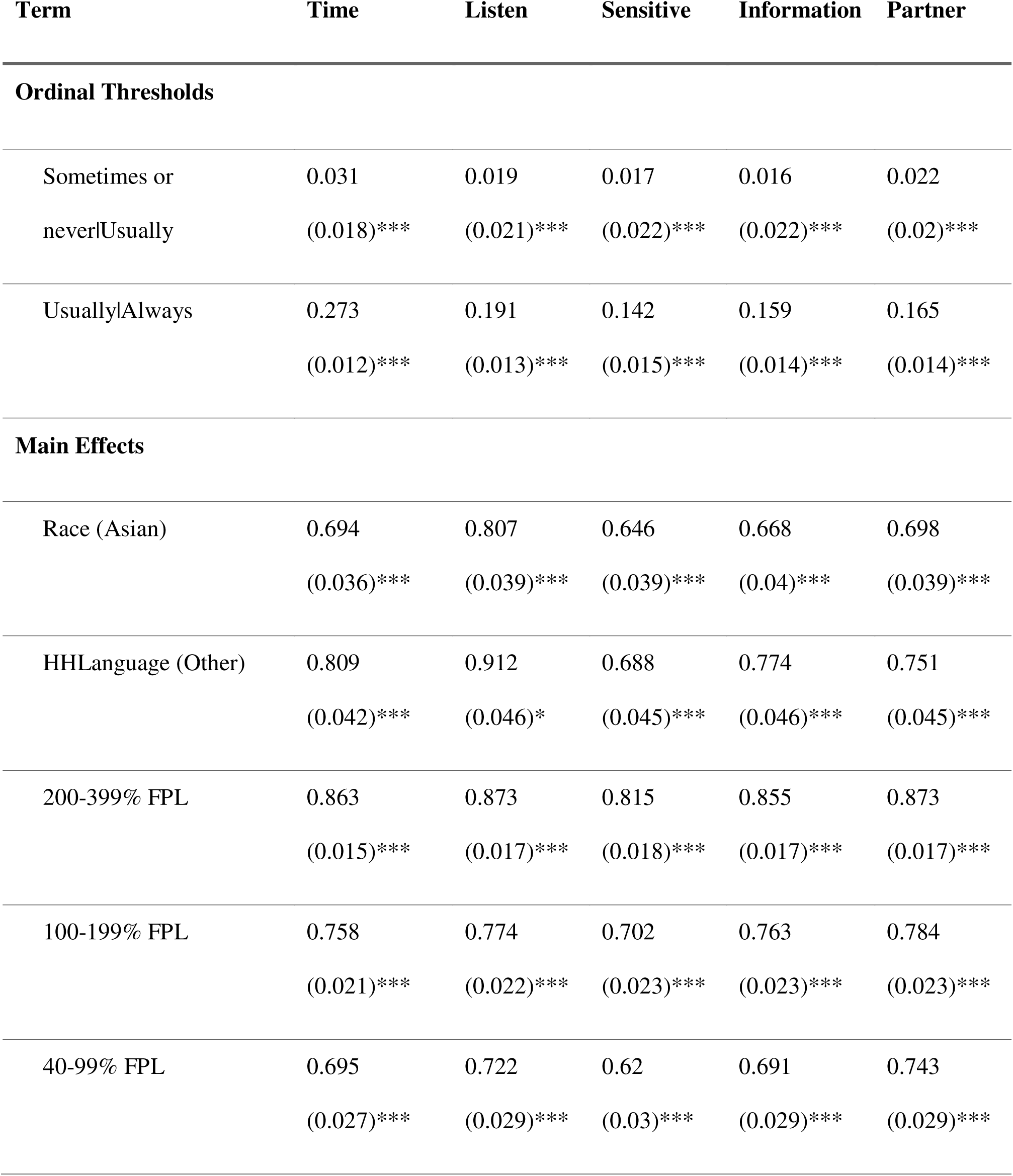
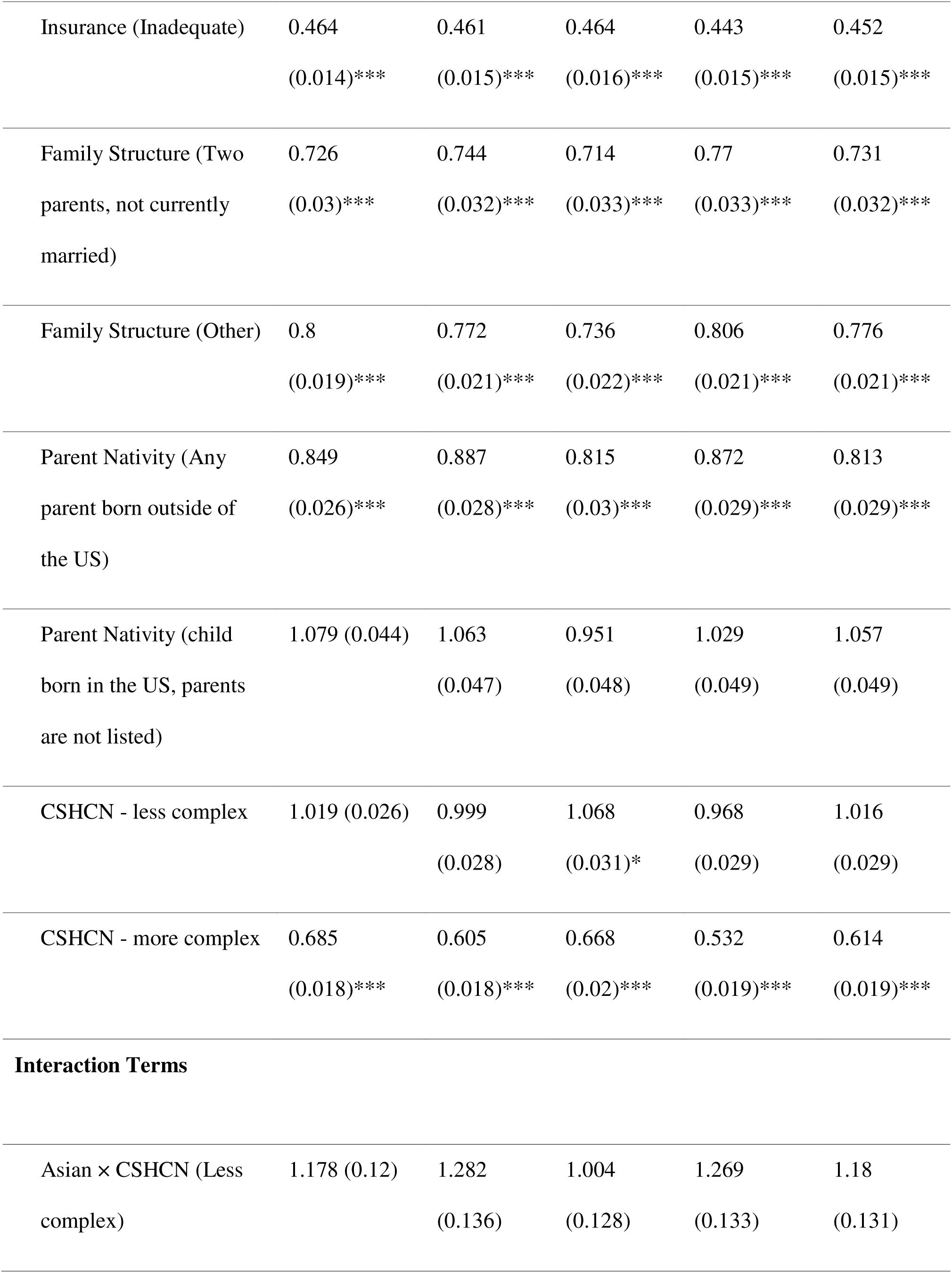
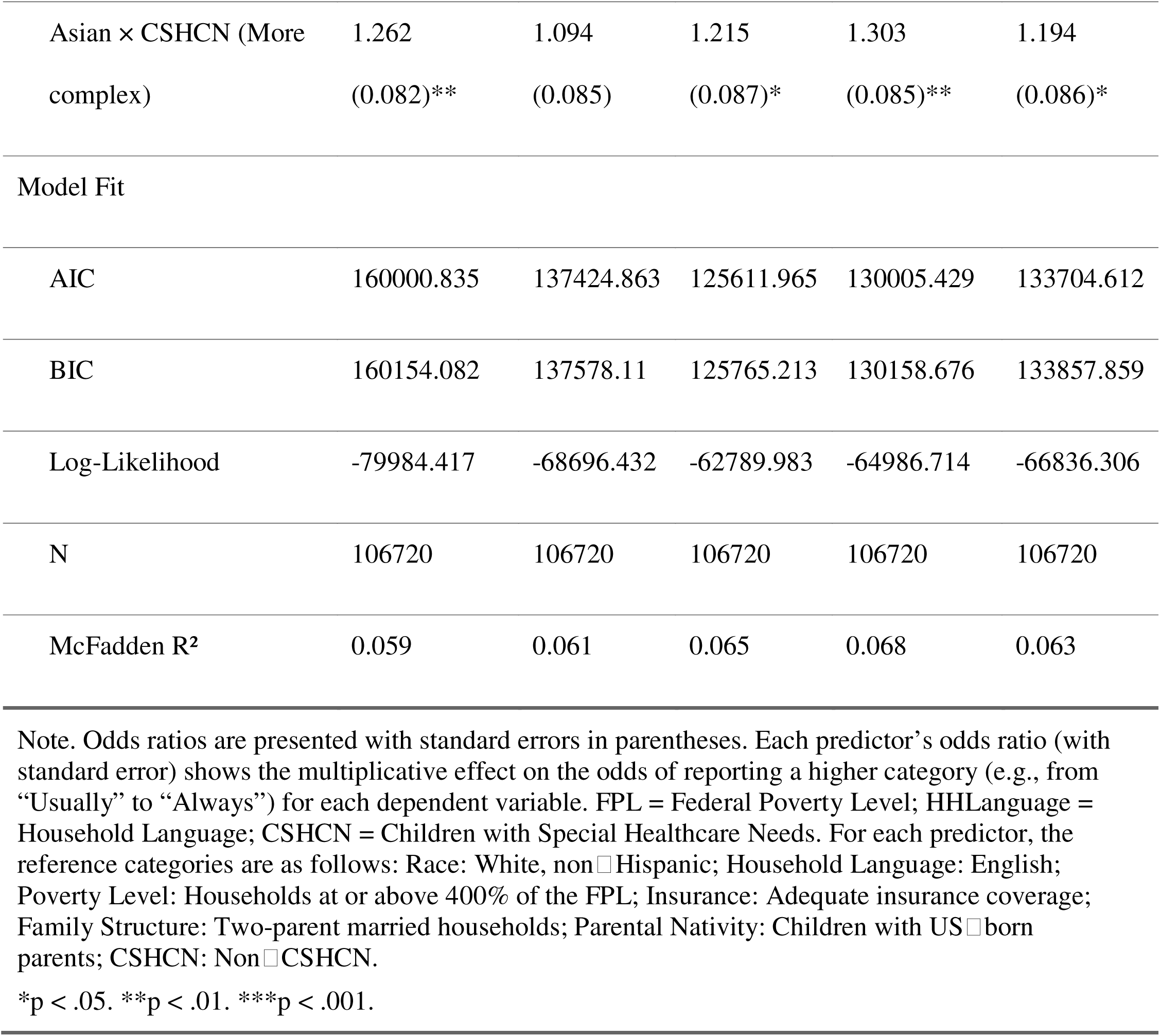
Multivariate Ordinal Regressions of Indicators of Healthcare Quality among Asian and Non-Hispanic White families (Odds Ratios)

| Term | Time | Listen | Sensitive | Information | Partner |
| --- | --- | --- | --- | --- | --- |
| **Ordinal Thresholds** |  |  |  |  |  |
| Sometimes or<br>never\|Usually | 0.031<br>(0.018)*** | 0.019<br>(0.021)*** | 0.017<br>(0.022)*** | 0.016<br>(0.022)*** | 0.022<br>(0.02)*** |
| Usually\|Always | 0.273<br>(0.012)*** | 0.191<br>(0.013)*** | 0.142<br>(0.015)*** | 0.159<br>(0.014)*** | 0.165<br>(0.014)*** |
| **Main Effects** |  |  |  |  |  |
| Race (Asian) | 0.694<br>(0.036)*** | 0.807<br>(0.039)*** | 0.646<br>(0.039)*** | 0.668<br>(0.04)*** | 0.698<br>(0.039)*** |
| HHLanguage (Other) | 0.809<br>(0.042)*** | 0.912<br>(0.046)* | 0.688<br>(0.045)*** | 0.774<br>(0.046)*** | 0.751<br>(0.045)*** |
| 200-399% FPL | 0.863<br>(0.015)*** | 0.873<br>(0.017)*** | 0.815<br>(0.018)*** | 0.855<br>(0.017)*** | 0.873<br>(0.017)*** |
| 100-199% FPL | 0.758<br>(0.021)*** | 0.774<br>(0.022)*** | 0.702<br>(0.023)*** | 0.763<br>(0.023)*** | 0.784<br>(0.023)*** |
| 40-99% FPL | 0.695<br>(0.027)*** | 0.722<br>(0.029)*** | 0.62<br>(0.03)*** | 0.691<br>(0.029)*** | 0.743<br>(0.029)*** |
| Insurance (Inadequate) | 0.464<br>(0.014)*** | 0.461<br>(0.015)*** | 0.464<br>(0.016)*** | 0.443<br>(0.015)*** | 0.452<br>(0.015)*** |
| Family Structure (Two<br>parents, not currently<br>married) | 0.726<br>(0.03)*** | 0.744<br>(0.032)*** | 0.714<br>(0.033)*** | 0.77<br>(0.033)*** | 0.731<br>(0.032)*** |
| Family Structure (Other) | 0.8<br>(0.019)*** | 0.772<br>(0.021)*** | 0.736<br>(0.022)*** | 0.806<br>(0.021)*** | 0.776<br>(0.021)*** |
| Parent Nativity (Any<br>parent born outside of<br>the US) | 0.849<br>(0.026)*** | 0.887<br>(0.028)*** | 0.815<br>(0.03)*** | 0.872<br>(0.029)*** | 0.813<br>(0.029)*** |
| Parent Nativity (child<br>born in the US, parents<br>are not listed) | 1.079 (0.044) | 1.063<br>(0.047) | 0.951<br>(0.048) | 1.029<br>(0.049) | 1.057<br>(0.049) |
| CSHCN - less complex | 1.019 (0.026) | 0.999<br>(0.028) | 1.068<br>(0.031)* | 0.968<br>(0.029) | 1.016<br>(0.029) |
| CSHCN - more complex | 0.685<br>(0.018)*** | 0.605<br>(0.018)*** | 0.668<br>(0.02)*** | 0.532<br>(0.019)*** | 0.614<br>(0.019)*** |
| **Interaction Terms** |  |  |  |  |  |
| Asian × CSHCN (Less<br>complex) | 1.178 (0.12) | 1.282<br>(0.136) | 1.004<br>(0.128) | 1.269<br>(0.133) | 1.18<br>(0.131) |
| Asian × CSHCN (More complex) | 1.262<br>(0.082)** | 1.094<br>(0.085) | 1.215<br>(0.087)* | 1.303<br>(0.085)** | 1.194<br>(0.086)* |
| Model Fit |  |  |  |  |  |
| AIC | 160000.835 | 137424.863 | 125611.965 | 130005.429 | 133704.612 |
| BIC | 160154.082 | 137578.11 | 125765.213 | 130158.676 | 133857.859 |
| Log-Likelihood | -79984.417 | -68696.432 | -62789.983 | -64986.714 | -66836.306 |
| N | 106720 | 106720 | 106720 | 106720 | 106720 |
| McFadden R<sup>2</sup> | 0.059 | 0.061 | 0.065 | 0.068 | 0.063 |

Note. Odds ratios are presented with standard errors in parentheses. Each predictor's odds ratio (with standard error) shows the multiplicative effect on the odds of reporting a higher category (e.g., from "Usually" to "Always") for each dependent variable. FPL = Federal Poverty Level; HHLanguage = Household Language; CSHCN = Children with Special Healthcare Needs. For each predictor, the reference categories are as follows: Race: White, non-Hispanic; Household Language: English; Poverty Level: Households at or above 400% of the FPL; Insurance: Adequate insurance coverage; Family Structure: Two-parent married households; Parental Nativity: Children with US-born parents; CSHCN: Non-CSHCN.
\*p < .05. \*\*p < .01. \*\*\*p < .001.

Race/Ethnicity emerged as a consistent predictor across all outcomes. As shown in Table 2, Asian, non-Hispanic parents were significantly less likely than White, non-Hispanic parents to report the highest level of perceived quality (e.g., “Always” time spent, “Always” listening). Odds ratios ranged from 0.65 to 0.81, indicating a substantively lower likelihood of optimal ratings for Asian families. In Figure 1, these differences are reflected in consistently lower predicted probabilities of “Always,” especially at the Non-CSHCN and Less Complex levels.

Household language was also associated with perceived quality, such that parents who spoke a language other than English at home had lower odds of reporting the highest quality ratings in several domains (ORs generally between 0.69 and 0.91). Poverty level demonstrated a clear dose–response relationship: families with lower incomes (e.g., 0–99% FPL) were significantly less likely to report optimal care experiences compared to families at or above 400% of the federal poverty level, with ORs in the 0.62–0.74 range across outcomes. Insurance adequacy was among the strongest predictors, as inadequate insurance was associated with substantially lower odds of optimal care (ORs ~ 0.44–0.47).

Family structure also influenced parents’ perceptions, with families headed by two unmarried parents or other family types having reduced odds of rating care as “Always” high quality (ORs ~ 0.71–0.80). Parental nativity revealed that having any parent born outside the US was associated with decreased odds of optimal ratings for several outcomes (ORs ~ 0.72–0.88). In contrast, the “Other” category of parental nativity (in which parents were not clearly classified) did not differ significantly from US-born parents for most outcomes.

CSHCN complexity showed a negative main effect, particularly at the “more complex” level. Families of children with more complex healthcare needs were markedly less likely to perceive optimal care across all domains (ORs ~ 0.53–0.69). However, as depicted in Figure 1, the interaction between Race/Ethnicity and CSHCN complexity suggested a partial attenuation of racial/ethnic disparities at higher levels of complexity. In particular, while both White and Asian families experienced lower predicted probabilities of “Always” ratings when healthcare needs were more complex, the gap between White and Asian families narrowed slightly. This pattern implies that the demands of more complex care may diminish, though not eliminate, some of the racial/ethnic differences seen at lower levels of complexity.

Overall, the models explained a modest but meaningful portion of the variance in parents’ perceptions of healthcare quality, as indicated by McFadden R² values ranging from 0.059 to 0.068.

## Discussion

This study sought to examine and raise awareness about the disparities in healthcare quality experienced by Asian CSHCNs. The examination specifically focused on how race, household language, poverty, family structure, and other elements affect the quality of care received by Asian and White CSHCNs. Results indicated notable findings. After controlling for other factors in the model, parents of Asian CSHCNs were overall significantly less likely to report high-quality care than parents of White CSHCNs in several areas, particularly in culturally sensitive care, obtaining information that was relevant to their concerns, doctors spending enough time with the child, and feeling like a partner in their child’s care. Additional and modifiable predictors were identified, including speaking another language, generational status, poverty, and insurance coverage. Families who reported that their household language was other than English, had inadequate insurance, had lower incomes (e.g., 0–99% FPL), led by two unmarried parents or other family types, and had any parent born outside the US were more inclined to reveal that they did not always receive high-quality care. Children with more complex special health care needs continue to experience lower quality of care than children with less complex special health care needs.

The interaction between race and CSHCN complexity was consistent across quality indicators. At the lowest quality rating (“sometimes or never”), differences in perceived care were minimal between Asian and White families as well as between families of children with and without special healthcare needs. However, at the intermediate (“usually”) level, Asian families reported slightly higher quality than White families, and families with more complex healthcare needs tended to report higher quality compared to those with less complex or no special needs. In contrast, at the highest quality rating (“always”), Asian families were less likely than White families to report that their child consistently received high-quality care, and families with more complex healthcare needs were also less likely to report always receiving high-quality care. The effect of race appears to be lessened at the highest level of CSHCN complexity.

These results were also comparable in various studies. Dembo et al. (2022) investigated the rate of determinants of racial and ethnic disparities in health and health care among CSHCN in Boston, Massachusetts. Although this study was limited due to various factors, it found notable results in racial health disparities (Dembo et al., 2022). Determinants, such as household income, early exposure to adverse childhood experiences, family structure, lack of neighborhood support, and household language, were significant factors in creating disparities between White, Black, and Hispanic CSHCNs (Dembo et al., 2022). Another study indicated similar results. Houtrow et al. (2011) aimed to identify and compare the overall health and service characteristics of CHSCN with and without disabilities and to determine factors that caused unmet need among these groups. Results found that decreased odds of unmet need were associated with families who were in or near poverty, low education levels in adults in the household, those living in single-mother households, and did not have adequate health insurance coverage (Houtrow et al., 2011). In addition to demonstrating parallel results regarding social determinants of health contributing to the increase of unmet need among families with CSHCN, they also found that CSHCN with disabilities (identified as CSHCN with a high level of severity and frequent occurrence of irregular health) had higher rates of need and unmet need when compared to other CSHCNs without disabilities (Houtrow et al., 2011). Areas of unmet need include specialty care, therapy services, mental health services, home health, assistive devices, medical supplies, and durable medical equipment (Houtrow et al., 2011). Although most studies concentrated on different races and included additional variables to assess determinants and health disparities, results still reflect the need to effectively improve access to and quality of healthcare delivered for minorities and CSHCN.

### Implications for Practice, Policy, and Research

First, healthcare providers should receive comprehensive training in cultural sensitivity and humility to better understand and respect the diverse backgrounds of Asian families. This includes recognizing cultural beliefs, values, and communication style that may influence care perceptions (Brooks et al., 2019). It is essential to prioritize a deeper understanding among healthcare providers regarding the factors that contribute to misunderstandings and uncomfortable interactions in cross-cultural communications with parents, particularly those from immigrant backgrounds (King et al, 2015). In the case of Asian parents, their communication style tends to be more indirect, and their decision-making processes often reflect a hierarchical structure, starkly contrasting with the direct and consensual communication commonly found in Western cultures. This disparity in cultural norms can create significant barriers; the authoritative role of healthcare providers may discourage parents from openly expressing their concerns. Therefore, it may hinder the development of a trusting relationship that fosters a family-professional partnership, recognizing parents as integral members of the healthcare team (Agency for Healthcare Research and Quality [AHRQ], 2022). Previous studies underscore the critical role of healthcare providers’ cultural awareness, active listening, empathy, and patience (Brooks et al., 2019; King, 2022); to actively engage families in care planning and decision-making processes. This inclusive approach involves constant attention, reflection, and action to understand the families’ perspectives and finding the most urgent needs, and it would help parents to be comfortable, informed and activated to understand and advocate for their child’s needs. For example, parents who are inadequately informed and activated about their child’s health condition may lack critical knowledge and motives to pursue preventative and specialty care that improves health outcomes (Eneriz-Wiemer et al., 2014; Son et al., 2018).

Second, implementing professional medical interpreter services and providing translated health materials can help bridge communication gaps for families with limited English proficiency. It is also essential to train healthcare providers on how to work effectively with interpreters. Enhancing interpretation services through pre-meetings, training, accommodations, and the use of peer navigators is crucial to meeting the needs of immigrant families. For example, it is essential to implement policies that establish comprehensive systems. These systems should include securing funding for the training of professionals and interpreters to enhance cross-cultural communication skills and deepen understanding of disabilities within healthcare settings. A notable gap exists in the preparedness of interpreters to effectively serve these communities. Establishing a structured approach that allows immigrant families to engage in pre-meetings with interpreters can be beneficial. Such meetings would provide an opportunity for families to articulate their specific needs and outline health plans for their children, enabling interpreters to prepare adequately for subsequent interactions. These preparatory sessions can foster a collaborative environment characterized by interdependent relationships, mutual trust, and shared responsibility between interpreters and families (Fukui et al., 2024; Hadziabdic et al., 2013). By doing so, interpreters can transition from mere translators to active partners in the care processes, ensuring that the services provided are culturally sensitive and tailored to the unique needs of Asian immigrant families.

Third, offering services such as transportation assistance, flexible scheduling, and patient navigation can alleviate barriers faced by low-income families, improving their access to consistent and high-quality care. In particular, implementing peer health navigator programs has proven effective in assisting immigrant parents in navigating complex healthcare systems. These programs have been adopted in the US and other countries to support families with limited English proficiency (Crezee, 2019; Grant et al., 2024; Shommu et al., 2016). A study by Crezee and Roat (2019) highlights the distinct roles of bilingual patient navigators compared to healthcare interpreters. While interpreters primarily focus on translating language, patient navigators engage more deeply by providing education, advocacy, and support tailored to families with low health literacy. Additionally, funding and supporting community health initiatives that engage with Asian communities can promote awareness of available services and encourage utilization of preventive and specialty care services among low-income Asian families with CSHCN.

Lastly, future studies should investigate the variations in healthcare experiences across different Asian subgroups, accounting for factors such as ethnicity, immigration status, and levels of acculturation. Furthermore, research should assess the effectiveness of cultural humility training programs on patient satisfaction and health outcomes within diverse populations. Longitudinal studies are also needed to understand how interventions targeting language and cultural barriers impact perceptions of healthcare quality over time, particularly among Asian families raising CSHCN.

Efforts to improve access to care for CSHCN are also needed. Buy-in from institutional leadership to provide abundant support is needed to alleviate the challenges and improve the quality of care received by CSHCN. In a study conducted in Ireland, researchers created a bespoke model to develop a team specifically focused on delivering high-quality clinical care for children with complex healthcare needs (Brenner et al., 2021). The bespoke model, designed for specific scenarios, was used to center methods on family-centered care (Brenner et al., 2021). Parents were encouraged to collaborate with service providers, serve as a knowledgeable resource to internal and external stakeholders, and supply coordinated and optimal services to children with complex healthcare needs (Brenner et al., 2021). Results found that establishing the service led to increased satisfaction from the parents with the improved quality of clinical care received and needed for their child (Brenner et al., 2021). Other strategies, such as hiring more advanced practice nurses and clinical nurse specialists to support communities and utilizing comprehensive and proactive care coordination strategies in medical homes for CSHCN, also demonstrated improvement in healthcare quality for children with complex special health care needs (Caicedo, 2016; Van Cleave et al., 2015).

## Conclusion

Addressing disparities in quality of care among Asian CSHCN requires a comprehensive strategy. Our findings show Asian parents report lower quality care than White parents, influenced by language barriers, socioeconomic factors, and cultural differences. Strategies to mitigate these disparities include provider training in cultural humility, improved language access, care coordination for low-income families, and the use of culturally matched peer health navigators to improve healthcare navigation and family empowerment. Future research should assess these interventions and explore subgroup differences to ensure equitable care.

## Data Availability

All data produced in the present study are available upon reasonable request to the authors.

## Glossary

CSHCN: Children with Special Healthcare Needs
FPL: Federal Poverty Level
HHLanguage: Household Language
NSCH: National Survey of Children’s Health

## Acknowledgements

We would like to thank the VCU Guided Research Experience and Applied Training (GREAT) Program for preparing undergraduate students to conduct responsible research, understand various topics in health and behavioral sciences, and present findings at local and national conferences. We would also like to thank Amy Adkins and Herbert Hill for their support, guidance, and dedication to mentoring students in furthering their research and goals.

## Author Contributions

**Kayla Van:** Conceptualization, data curation, writing – original draft, review, and editing

**Chin-Chih Chen:** Conceptualization, writing – original draft, review, and editing

**Esther Son:** Conceptualization, writing – original draft, review, and editing

**Ruth Brown:** Conceptualization, writing – original draft, review, and editing, data analysis, supervision

